# Randomized clinical trial quality has improved over time but is still not good enough: an analysis of 176,620 randomized controlled trials published between 1966 and 2018

**DOI:** 10.1101/2020.04.22.20072371

**Authors:** Christiaan H. Vinkers, Herm J. Lamberink, Joeri K. Tijdink, Pauline Heus, Lex Bouter, Paul Glasziou, David Moher, Johanna A. Damen, Lotty Hooft, Willem M. Otte

**Author notes:** Corresponding author Christiaan H. Vinkers MD, Department of Psychiatry, Amsterdam University Medical Center (UMC), De Boelelaan 1108 (Room 13E11), 1081 HZ Amsterdam, the Netherlands.

## Abstract

**Background:** Many randomized controlled trials (RCTs) are biased and difficult to reproduce due to methodological flaws and poor reporting. There is increasing attention for responsible research practices including reporting guidelines, but it is unknown whether these efforts have improved RCT quality (i.e. reduced risk of bias). We therefore mapped trends over time in trial publication, trial registration, reporting according to CONSORT, and characteristics of publication and authors.

**Methods:** Meta-information of 176,620 RCTs published between 1966 and 2018 was extracted. Risk of bias probability (four domains: random sequence generation, allocation concealment, blinding of patients/personnel, and blinding of outcome assessment) was assessed using validated risk-of-bias machine learning tools. In addition, trial registration and reporting according to CONSORT were assessed with automated searches. Characteristics were extracted related to publication (number of authors, journal impact factor, medical discipline) and authors (gender and Hirsch-index).

**Findings:** The annual number of published RCTs substantially increased over four decades, accompanied by increases in the number of authors (5.2 to 7.8), institutions (2.9 to 4.8), female authors (20 to 42%, first authorship; 17 to 29%, last authorship), and Hirsch-indices (10 to 14, first authorship; 16 to 28, last authorship). Risk of bias remained present in most RCTs but decreased over time for the domains allocation concealment (63 to 51%), random sequence generation (57 to 36%), and blinding of outcome assessment (58 to 52%). Trial registration (37 to 47%) and CONSORT (1 to 20%) rapidly increased in the latest period. In journals with higher impact factor (>10), risk of bias was consistently lower, higher levels of trial registration more frequent, and mentioning CONSORT.

**Interpretation:** The likelihood of bias in RCTs has generally decreased over the last decades. This may be driven by increased knowledge and improved education, augmented by mandatory trial registration, and more stringent reporting guidelines and journal requirements. Nevertheless, relatively high probabilities of bias remain, particularly in journals with lower impact factors. This emphasizes that further improvement of RCT registration, conduct, and reporting is still urgently needed.

**Funding:** This study was funded by The Netherlands Organisation for Health Research and Development (445001002).

## Introduction

Randomized controlled trials (RCTs) are the primary source for evidence on the efficacy and safety of clinical interventions, and systematic reviews and clinical guidelines synthesize their results. Unfortunately, many RCTs have severe methodological flaws and results are often biased.^1^ Strikingly, the majority of RCT findings have inflated estimates and have problems with randomization, allocation concealment, and blinding.^2,3^ Recently, it was shown that over 40% of RCTs were at high risk of bias which could have been easily avoided.^4^ Moreover, poor reporting prevents the adequate assessment of RCT quality and limits its reproducibility.^5^ Avoidable sources of waste and inefficiency in clinical research were estimated to be as high as 85%.^6^

For a longer time period, CONSORT criteria have been introduced to improve RCT reporting, and mandatory RCT registration by the International Committee of Medical Journal Editors (ICMJE) has been put forward,^7^.^8^ More recently, *The Lancet* published a series on increasing value and reducing waste in medical research which proposed meaningful steps towards more high-quality research, including improved methodology and reporting, and reduction of unpublished negative findings.^5,9^ Additional actions to improve RCT quality and transparency include trial tracker initiatives aimed at reducing non-publication of clinical trials,^10^ and fostering responsible research practices. At the most recent World Conference on Research Integrity, the Hong Kong Principles were proposed to further stimulate responsible research practices by including them in researcher assessments.^11^

Even though these actions and initiatives have undoubtedly contributed to awareness that the quality of RCTs needs to improve, the question remains whether real progress has been made in reducing the extent of avoidable waste in clinical research. In other words, have these initiatives and measures improved the quality, transparency, and reproducibility of RCTs? Several studies have assessed the quality of reporting and risk of bias in RCTs,^12^ but most are relatively small and limited to specific medical disciplines or time periods. Nevertheless, based on 20,920 RCTs from Cochrane reviews, there are indications that poor reporting and inadequate methods have decreased over time.^13^ However, large-scale evidence on trends of RCT characteristics and quality across medical disciplines over time is currently lacking. This is surprising in view of the importance of valid and reliable evidence from RCTs for patient care. Therefore, this study aimed to provide a comprehensive and unbiased analysis of developments in the clinical trial landscape between 1966 and 2018 based on 176,620 full-text RCT publications.

## Methods

The protocol for this analysis was registered prior to study conduct,^14^ the database and scripts are available through GitHub (see Data Sharing), and the results are disseminated through the medRxiv pre-print server.

### Selection of RCTs and extraction of characteristics

RCTs were identified via MEDLINE (Nov 20, 2017) starting with all publications indicated as ‘randomized controlled trial’ using the query “randomized controlled trial[pt] NOT (animals[mh] NOT humans[mh])”. The initial search did not include a time window. Non-English language, non-randomized, animal, pilot, and feasibility studies were subsequently excluded (see **Supplementary Methods** text for details on selection procedure). We collected the Portable Document Format (PDF) for all available RCTs across publishers in journals covered by the library subscription of our institution, and converted these PDFs to structured text in xml format using publicly available software (Grobid, available via GitHub). By linking information from the MEDLINE database, the full-text publication, and data from Scopus and Web of Science, we extracted metadata on number of authors, author gender, number of countries and institutions of (co-)authors, and the Hirsch (H)-index of the first and last authors (see **Supplementary Table S1** for details). Moreover, we extracted the journal impact factor (JIF) at the time of publication. We also quantified the frequency of predefined positive, negative, and neutral words (25 words in each category) in titles and abstract texts as previously published (see **Supplementary Methods** for details).^15^ Time was stratified in 5-year periods as behavioral changes are expected to occur with relative low pace, with the relatively few trials published before 1990 merged in one stratum.

### Risk of Bias assessment

For every included full-text RCT, risk of bias assessment was automatically performed using machine learning assessment developed by RobotReviewer.^16^ This tool is optimized for large-scale characterizations^17,18^ and algorithmically based on a large sample of human-rated risk of bias reports and extracted support texts from trial publications covering the full RCT spectrum.

The level of agreement between RobotReviewer and human raters was similar for most domains (human-human agreement: min/max, 71-85%, average, 79%), human-RobotReviewer agreement: min/max 39-91%, average 65%).^20,21^ Of the seven risk of bias domains described by Cochrane,^19^ we assessed four: random sequence generation and allocation concealment (i.e., selection bias)), blinding of participants and personnel (i.e., performance bias), and blinding of outcome assessment (i.e., detection bias). Publication bias and outcome reporting bias were outside the scope of our analysis.

### Analysis of trial registration and CONSORT statement use

To check for trial registration, we extracted trial registration numbers from the abstract and full text publication and searched for the corresponding trial registration number in two online databases: WHO’s International Clinical Trials Registry Platform, composed of worldwide primary and partner registries, and the ClinicalTrials.gov trial registry.^20^ We checked all full-text publications for at least one mention of the words “Consolidated Standards of Reporting Trials” or CONSORT.

### Analysis related to Journal Impact Factor

Even though the journal impact factor (JIF, average number of times its articles has been cited in other articles for two years) is not a very suitable indicator of journal quality,^21^ no unbiased alternatives exist. In our study, we therefore used the JIF as a proxy to identify journals with high publication standards and high rejection rates. For each individual trial we selected the JIF of the year before trial publication. We used a JIF threshold of 10 as the primary cutoff based on JIF distributions (see **Supplementary Table S2** and **Supplementary Figure S1**) and previous evidence for sensitivity to assess RCT quality using this cutoff.^13^ However, we also performed sensitivity analyses for JIF cutoff thresholds at 3 and 5.

### Analyses related to medical disciplines

We assigned RCTs to medical disciplines based on the journal category (Web of Science).^7^ As a secondary analysis, we examined medical disciplines separately.^9^ Medical disciplines with less than 4,000 RCTs in our sample were assigned to the category ‘Other’.

### Power calculation and statistics

No a priori formal power calculation was performed as the aim of this project was to include all RCTs available on PubMed. Temporal patterns in the individual risk of bias scores were modeled with regression analysis. Reported P-values correspond to the comparison of the average value per year in the 1990–1995 and 2010–2018 strata. This post-1990 period was chosen to cover the first years following significant awareness on the need to report transparently, in comparison to the latest years in our dataset. Temporal patterns in trial registration and CONSORT use were modelled with logistic regression. Because median values were very close to mean values, the data is presented as mean ± 95% confidence intervals.

## Results

### RCT full-text acquisition process

From the 445,159 PubMed entries for RCTs, we identified 306,737 eligible RCTs (see flow chart in **Figure 1**). Full-text articles were obtained of 183,927 RCTs. RCT publications with an uncertain year of publication (7,307) were excluded, resulting in a final sample size of 176,620 RCTs.

**Figure 1:**
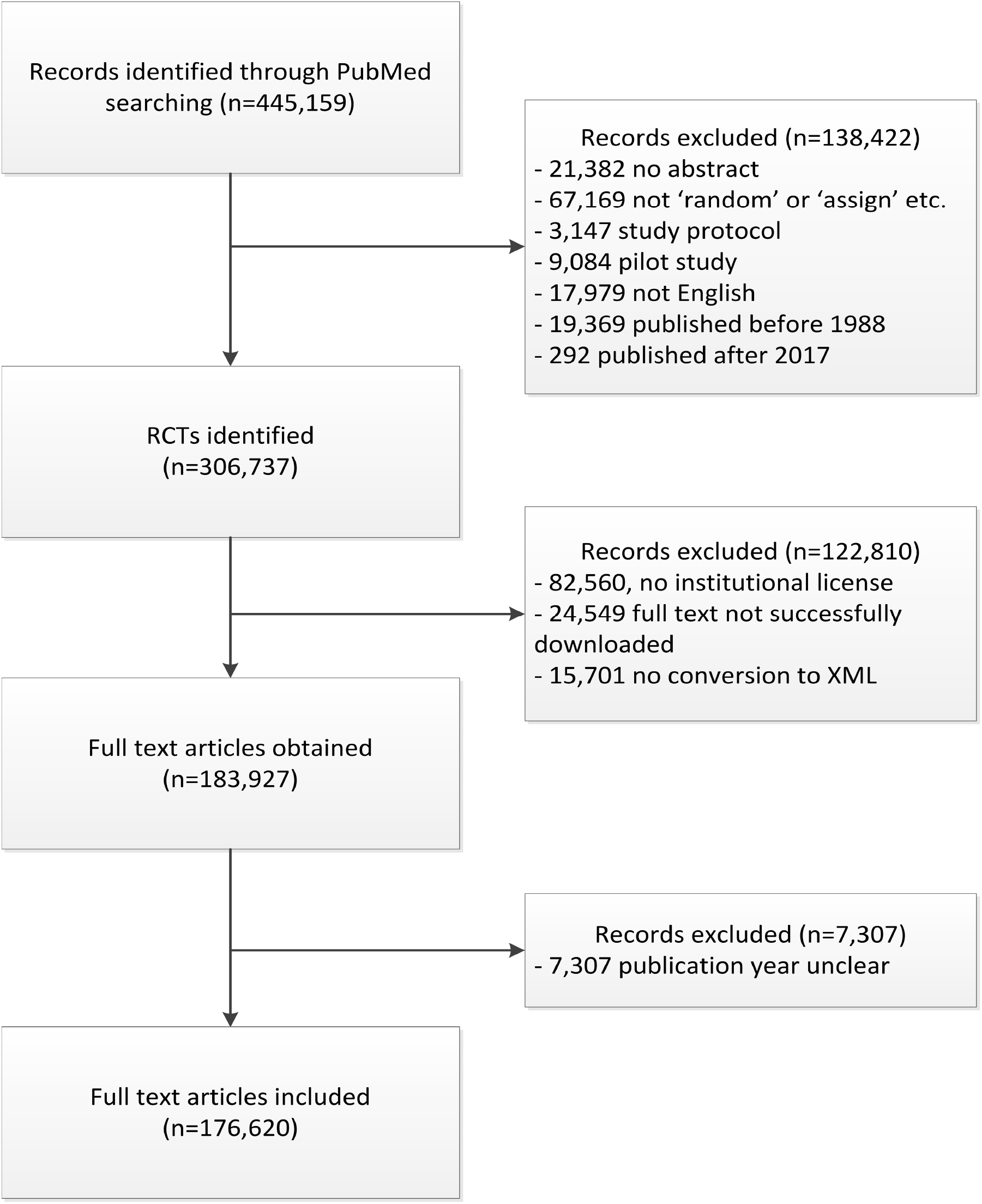
Flow chart of how the final full-text randomized clinical trials were obtained.

### RCT characteristics over time

RCTs for which full texts were obtained were predominantly published over the last three annual strata with 89,373 publications in the period 2010–2018 (11,172 per year) compared to 6,066 publications between 1990 and 1995 (1,213 per year; **Figure 2A**). Over time, the average number of authors steadily increased from 5.2 (CI: 5.12–5.26) in 1990–1995 to 7.8 in 2010– 2018 (CI: 7.76–7.83; P<0.0001) (**Figure 2B**). The percentage of female authors has gradually increased over time as well (**Figure 2C)**. As an example, the proportion of first and last female authors almost doubled from 20.4% (CI: 19.0–21.8%) in 1990–1995 to 42.4% (CI: 42.0–42.7%; P<0.001) in 2010–2018 for first female authors and from 17.4% (CI: 16.0–18.7%) in 1990– 1995 to 29.2% (CI: 28.9–29.5%; P<0.001) in 2010–2018 for last authors. The H-index of first and last authors of RCT publications also substantially increased over time from 9.9 (CI: 9.5– 10.2) in 1990–1995 to 14.4 (CI: 14.3–14.5; P<0.001) in 2010–2018 (**Figure 2D-E**). This was accompanied by a steady increase in the number of involved countries and institutions affiliated with all authors (number of institutions in 1990–1995: 3.24 (CI: 3.17–3.31) vs. 2010–2018: 4.84 (CI: 4.81–4.87; P<0.0001)) (**Figure 2F-G**). The frequency of negative words was stable over time, whereas the frequency of positive words significantly increased from 0.035% (0.031–0.040%; in 1990–1995), to 0.058% (0.056–0.059% in 2010–2018) (**Figure 2H**).

**Figure 2.**
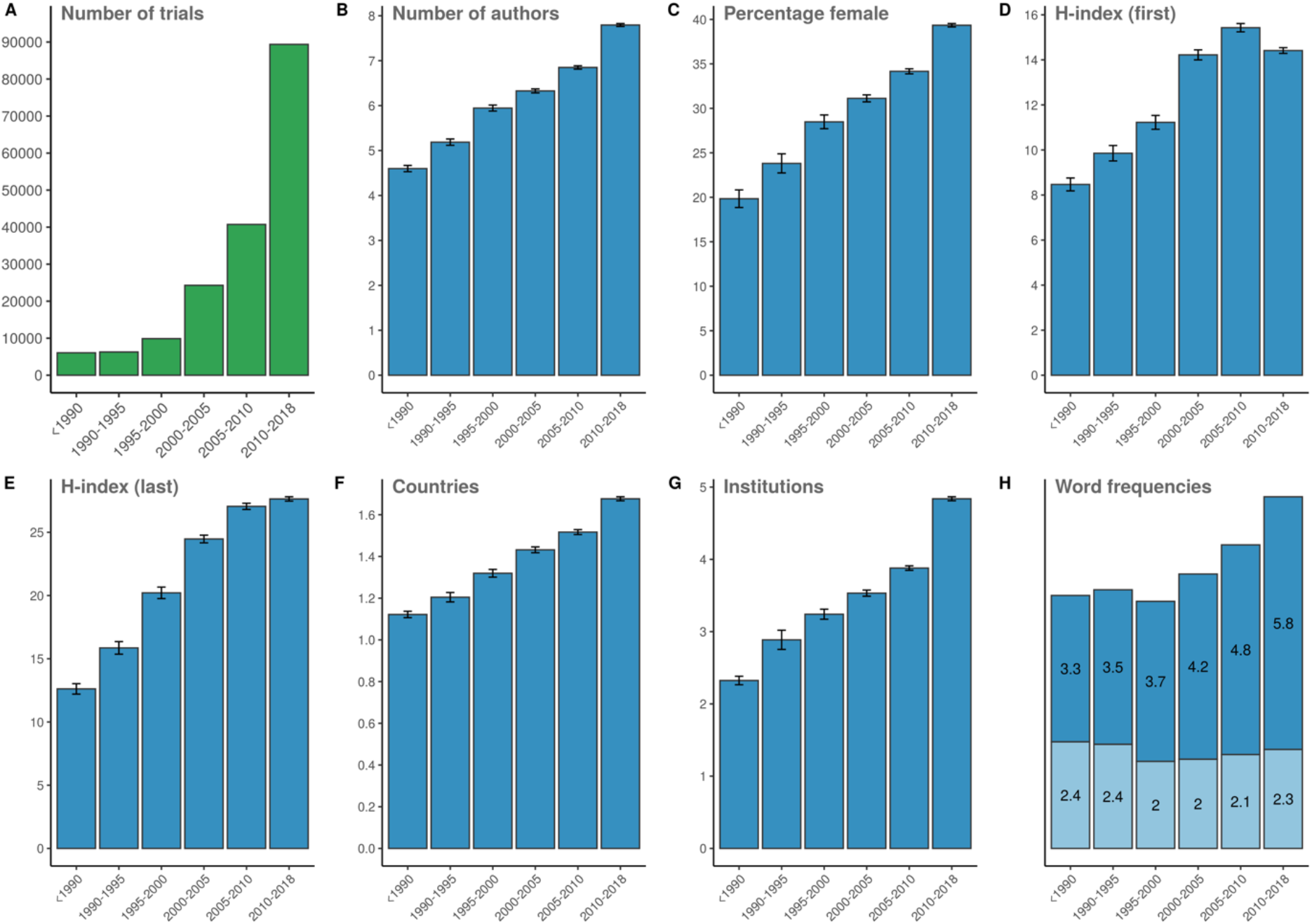
Number of RCTs included over time (A) and the corresponding average number of authors (B), percentage of female authors (C), H-index of the first author (D), H-index of the last author (E), number of countries involved (F), number of institutions involved (G), and the frequency of negative words (lower bars) and positive words (upper bars) in the RCT abstracts (H). Indicated stratum range is up to but not including the last year.

### Risk of bias, registration, and reporting: trends over time

We found an overall continuous reduction in the risk of bias due to inadequate allocation concealment, dropping from 62.8% (CI: 62.4–63.1%) in trials published in 1990–1995 to 50.9% (CI: 50.7–51.0%; P<0.0001) in trials published between 2010 and 2018 (**Figure 3A**). There was a relatively stronger decrease in the risk of bias due to non-random sequence generation, from 54.0% (CI: 53.6–54.5%) for trials in 1990–1995 to 36.4% (36.3–36.6%; P<0.001) for trials in 2010–2018 (**Figure 3B**). The risk of bias due to not blinding participants and personnel showed a distinctly different pattern, with a sequential increase since 2000 up to 56.9 (56.8– 57.1%; P<0.001) in 2010–2018, after an initial decrease (**Figure 3C**). The risk of bias due to not blinding outcome assessment decreased over time, from 56.6 (56.4–56.9%) to 51.8 (51.7– 51.9%; P< 0.001; **Figure 3D**). In all RCTs, mention of a trial registration number rapidly increased from close to zero to up to 46.7% (46.4–47.0%) in 2010–2018 (**Figure 3E**). We found very low use (1.05 (CI: 0.82–1.35%) in 1990–1995) of the CONSORT Statement in full text RCT publications, contrasting with 19.5% (19.3–19.8%; P<0.0001) of all trials between 2010 and 2018 (**Figure 3F**).

**Figure 3.**
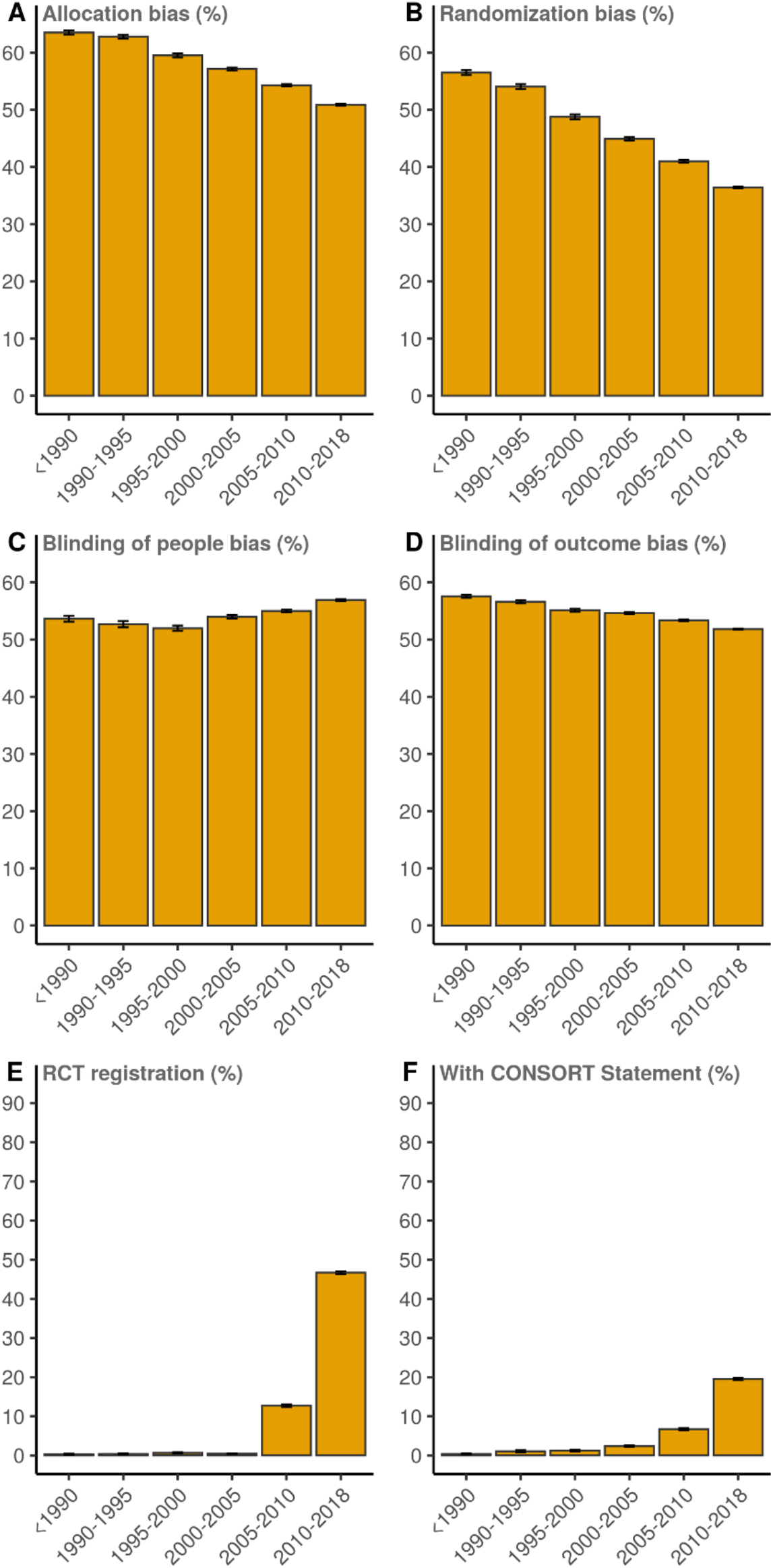
Risk of bias due to inadequate allocation concealment (A), random sequence generation bias (B), bias in blinding of patients and personnel (people) (C), bias in blinding of outcome assessment (D), RCT registration (E,) and mentioning of the CONSORT Statement (F) for all RCTs plotted over time. Indicated stratum range is up to but not including the last year.

### Risk of bias and reporting: relation with journal impact factor

The risk of bias in allocation concealment was consistently lower in trials published in journals with JIF larger than 10 (P< 0.001; **Figure 4A**). This also applied to randomization and blinding of participants and personnel and outcome assessment, even though the results were less pronounced compared to allocation concealment bias (P<0.0001, all domains, for latest time point; **Figure 4B-D**). Large differences were found in terms of trial registration and mentioning of CONSORT between RCTs published in high or lower impact journals (P<0.0001 for latest time point; **Figure 4E-F**). Seventy-three percent (72–74%) of trials in journals with a JIF higher than 10 was registered, and 26% (25–27%) mentioned the CONSORT Statement between 2010 and 2018 (both measures: P<0.0001 in comparison with JIF <10). Sensitivity analysis with JIF cutoff values of 3 and 5, respectively yielded comparable but smaller differences with reduced bias and increased registration and mentioning of the CONSORT Statement in journals with higher JIF (**Supplementary Figures S2 and S3**).

**Figure 4.**
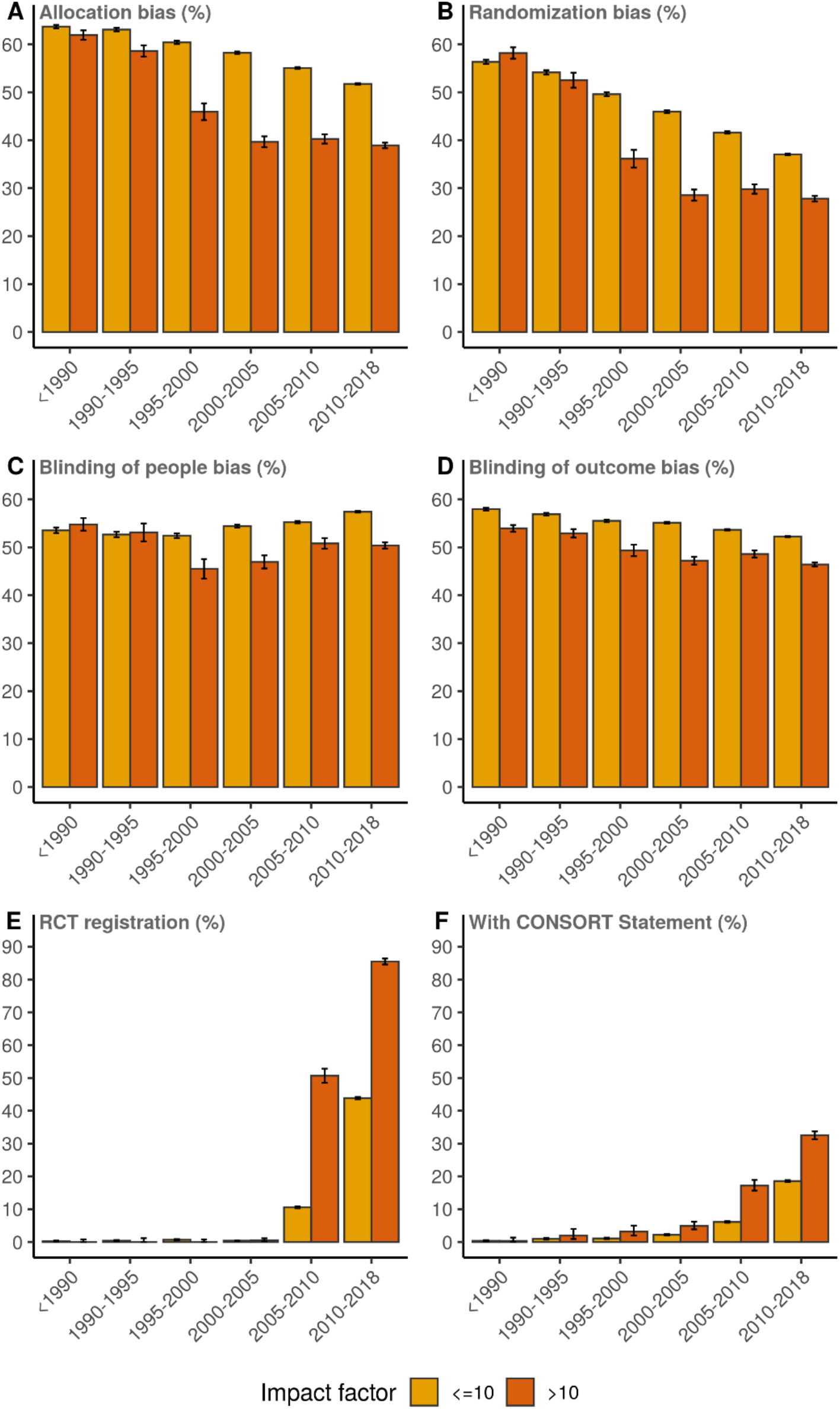
Risk of bias in allocation concealment (A), bias in randomization (B), bias in blinding of patients and personnel (people) (C), bias in blinding of outcome assessment (D), RCT registration (E) and mentioning of the CONSORT Statement (F) plotted over time plotted over time for RCTs published in journals with JIF>10 and journals with JIF<10. Indicated stratum range is up to but not including the last year.

### Risk of bias and reporting: relation with medical discipline

Risk of bias patterns substantially differed across medical disciplines (**Supplementary Table S1**). Lowest probabilities of bias were found in RCTs within the field of anesthesiology (27% randomization bias, 43% allocation concealment bias, 45% risk of bias due to insufficient blinding of participants and personnel, 45% bias in blinding of outcome assessment) (**Supplementary Figure S4**). The field of oncology had the highest levels of trial registration (43.4%) and mention of the CONSORT Statement (30.3%) (**Supplementary Figure S5**). Registration rates were lowest in the field of Endocrinology & metabolism (8.0%) and Urology & nephrology (10.2%).

## Discussion

We analyzed a total of 176,620 full-text publications of RCTs from the last four decades and show that the landscape of RCTs has considerably changed. Not only there is a growing number of authors and institutions from a growing number of countries involved in publications, but also a steadily increasing average H-index of the first and last author. The absolute and relative numbers of female authors in RCTs also gradually increased over time, with a large rise in first and last female authorships. Even though trends are increasing over time, the average percentage of female last authorships remains relatively low at 29% (2010–2018), in line with recent literature.^22^

With regard to risk of bias, the trends that emerge from our analyses are certainly hopeful. Risk of bias in RCTs declined over the past decades, with lowering trends for bias related to random sequence generation, allocation concealment and blinding of outcome assessment. In accordance, there is an increasing percentage of RCTs that are registered in public trial registers and in the use CONSORT guidelines. Despite two decades of documentation and calls for trial registration, it only substantially increased around 2004 when trial registration was made a condition for publication by International Committee of Medical Journal Editors (ICMJE). This policy was implemented and supported by the World Health Organization (WHO) in July 2005.^23^ Our results are in line with the assessment of 20,920 RCTs from Cochrane reviews in 2017 that found improvements in reporting and methods over time for sequence generation and allocation concealment.^13^

Notwithstanding these improvements, it is also clear that there is still a pressing need to further improve the quality of RCTs. The average risk in each of the bias domains remains generally high (around 50%), and bias related to blinding of participants and personnel is increasing over time, which may be due to more pragmatic or non-drug RCTs being performed. Moreover, despite the requirement of trial registration for publication since 2004, still in 2017 a substantial percentage of published RCTs are not registered. Furthermore, many RCTs do not mention the CONSORT guidelines in their full text, and more so for journals with lower impact factors. Despite accessibility of reporting guidelines, researchers are generally not required to adhere to them, and, more problematic, requirements are not strictly enforced and non-compliance to all the items on the reporting guideline is not sanctioned.^4,24^ To further improve the quality and reliability of RCTs, there is still a long way to go, and the rather slow progress of improvement may be due to the complex nature of conducting RCTs. Better education, enforcements, and (dis)incentives may be inevitable. Additionally, making data sets available according to the FAIR principles arguably will improve the situation.^25^

Depending on one’s expectations and future goals, the interpretation can be either optimistic or pessimistic: optimistic because, over the past decades, there has been quite some improvement in RCT conduct and reporting, but pessimistic because the improvements are going at a rather slow pace. From our analyses, it also appears that journals with higher JIF generally publish RCTs with lower scores on risk of bias domains. Our results confirm previous results showing higher JIF (higher than 10) being associated with a lower proportion of trials at unclear or high risk of bias in Cochrane reviews.^13^ Even though JIFs are not a very suitable measure of journal quality, our results are in line with previous studies showing that increased JIF is related with higher RCT quality.^26^ Finally, there are large differences across medical disciplines related to risk of bias scores across domains which cannot readily be explained.

There are several strengths and limitations inherent to our approach of automated extraction of full-text RCT publications. The automated and uniform approach yielded an unprecedented large and rich data source concerning RCTs from the last forty years is available for further study (see https://github.com/wmotte/frrp for the data), covering a large proportion of all published RCTs included in PubMed. Nevertheless, there are several limitations. First, risk of bias is inherently difficult to assess. Experts’ assessments of trials show that labeling the same trials for different Cochrane reviews resulted in substantial differences.^27,28^ Probabilities assigned with machine learning are based on a large set of human-assigned labels, and a direct comparison shows computerized assessment performance of 71.0% agreement.^29^ Second, we did not investigate all aspects of methodological rigor. In our study, we did not check for forms of attrition bias (e.g., incomplete outcome data) or reporting bias (e.g., selective outcome reporting), which both would require a direct comparison between the trial registration and actual trial publication report. Third, even though the CONSORT Statement was introduced to improve RCT reporting,^30^ the rapid increase of RCTs that mention following the CONSORT guideline does not guarantee adherence and reporting quality can remain suboptimal.^12^ We were not able to automatically correct for the conventional and non-abbreviated use of the word ‘consort’. This may have slightly increased our CONSORT Statement percentages and explains the very low but non-zero values in the earliest stratum.

Our comprehensive picture of RCT quality provides quantitative insight into the current state and trends over time. With many thousands of RCTs being published each year and thousands of clinical trials currently recruiting patients, this can help us to better understand the current situation but also to find solutions for further improvement. These could include a more stringent adoption of measures to enforce transparent and credible trial publication, but also fine-tuning of stricter registration regulations. In conclusion, our comprehensive analyses of a large body of full-text RCTs show that there is a slow and gradual improvement of RCT quality over the last decades. While RCTs certainly face challenges in relation to their quality and reproducibility and there is still ample room for improvement, our study is a first step in showing that all efforts that have been made to improve RCT practices may be paying off.

## Data sharing

The risk of bias characterization was done with a large-batch-customized-customized Python scripts (version 3; https://github.com/wmotte/robotreviewer_prob). The data management and analyses used R (version 3.6.1). All data including code and risk of bias data are available at https://github.com/wmotte/frrp).

## Data Availability

All data referred to in the manuscript is freely available: 10.5281/zenodo.3695150

https://zenodo.org/record/3695150#.Xp2KwFMzZDU

https://github.com/wmotte/frrp

## Supplementary Material

### Supplementary Methods

#### Data collection procedures

<u>Step 1. Identification of all human randomized controlled trials (RCTs)</u>. The Entrez API enables access to the PubMed database and was used via R Statistical Software using the query: “randomized controlled trial[pt] NOT (animals[mh] NOT humans[mh])”. Basic information (including authors, affiliations, journal, trial registry numbers, language, and funding agency) which is indexed by the PubMed database was downloaded.

<u>Step 2. Filtering identified studies.</u> Not all automatically identified studies were RCTs – despite the query in Step 1. To exclude potential contamination of the data by non-randomized, pilot and feasibility studies, a more strict selection of studies was made based on the title and abstract. Articles were automatically excluded in the following conditions:

- When the title contained: “study protocol”, “study design”, “protocol for”, “pilot” or “feasibility”
- When the abstract contained: “pilot study” or “feasibility study”
- When the title or abstract did not contain: “random” (in title and abstract) OR “assign” OR “allocat*” OR “placebo” OR “double-blind” (in abstract).
- When language was other than English

<u>Step 3. Downloading PDFs</u> of the remaining studies after Step 1 and 2. We used R scripts to download the PDF of each publication via the website of the respective publisher (https://github.com/wmotte/frrp). All downloaded PDFs were transformed to text data in the Extensible Markup Format (XML), using the open source software GROBID (https://github.com/kermitt2/grobid).

<u>Step 4. Retrieving additional data</u>. Using the PubMed identifier (PMID), we linked the included RCTs to databases such as the Cochrane Database of Systematic Reviews (CDSR) and Scopus, where we downloaded relevant information if a PMID is provided.

#### Positive and negative words used in title and abstract texts as previous published^15^

Positive words (n=25): Amazing, assuring, astonishing, bright, creative, encouraging, enormous, excellent, favourable, groundbreaking, hopeful, innovative, inspiring, inventive, novel, phenomenal, prominent, promising, reassuring, remarkable, robust, spectacular, supportive, unique, unprecedented

Negative words (n=25): Detrimental, disappointing, disconcerting, discouraging, disheartening, disturbing, frustrating, futile, hopeless, impossible, inadequate, ineffective, insignificant, insufficient, irrelevant, mediocre, pessimistic, substandard, unacceptable, unpromising, unsatisfactory, unsatisfying, useless, weak, worrisome

---

**Supplementary Figure S1.**
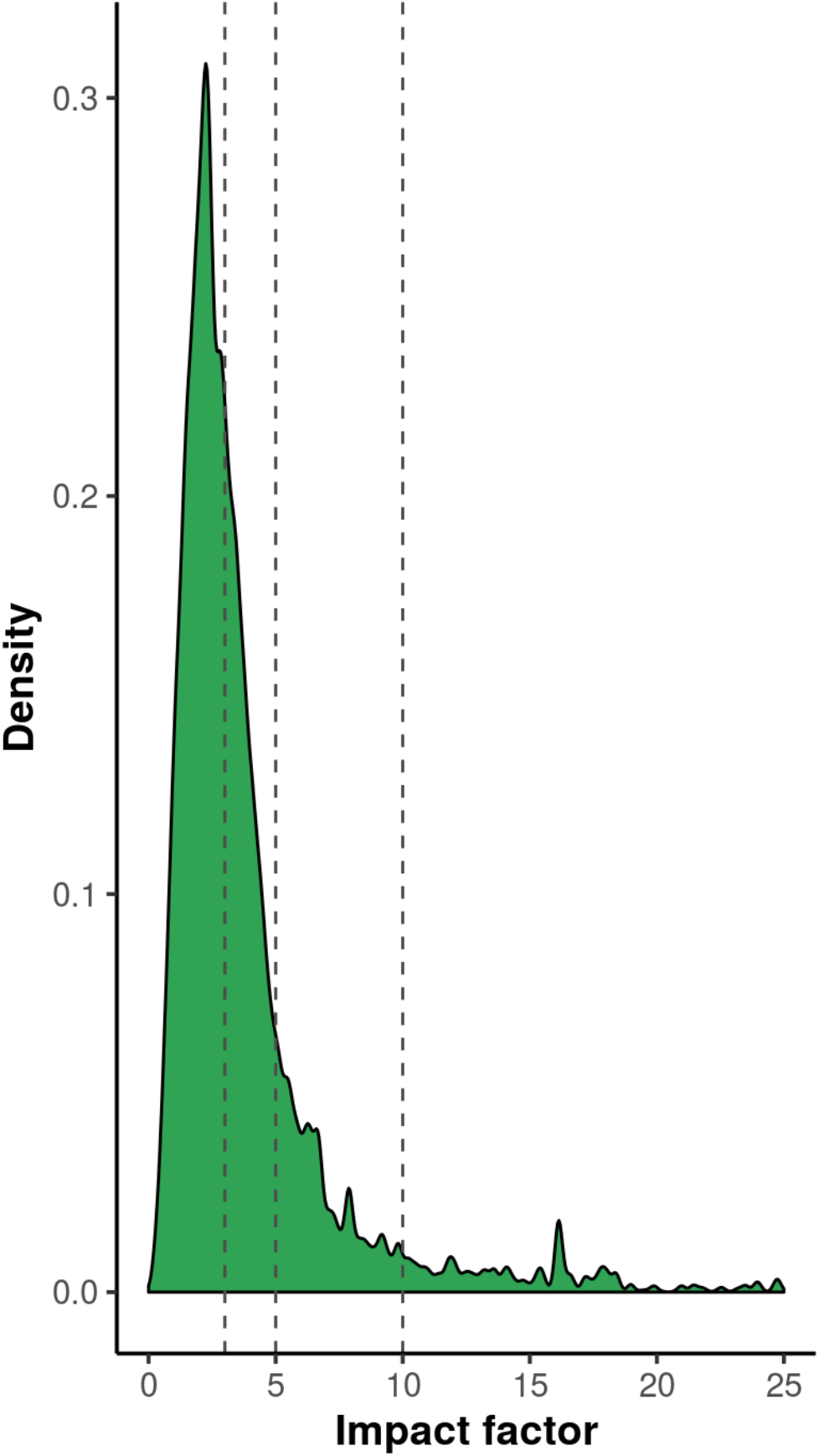
Distribution of journal impact factors (JIFs) of analyzed RCTs with JIF cutoffs of 3, 5 and 10 (dotted lines). The JIF of a journal in the year following the publication data of the RCT was used. Density represents the probability of a trial to belong to a given impact factor.

**Supplementary Figure S2.**
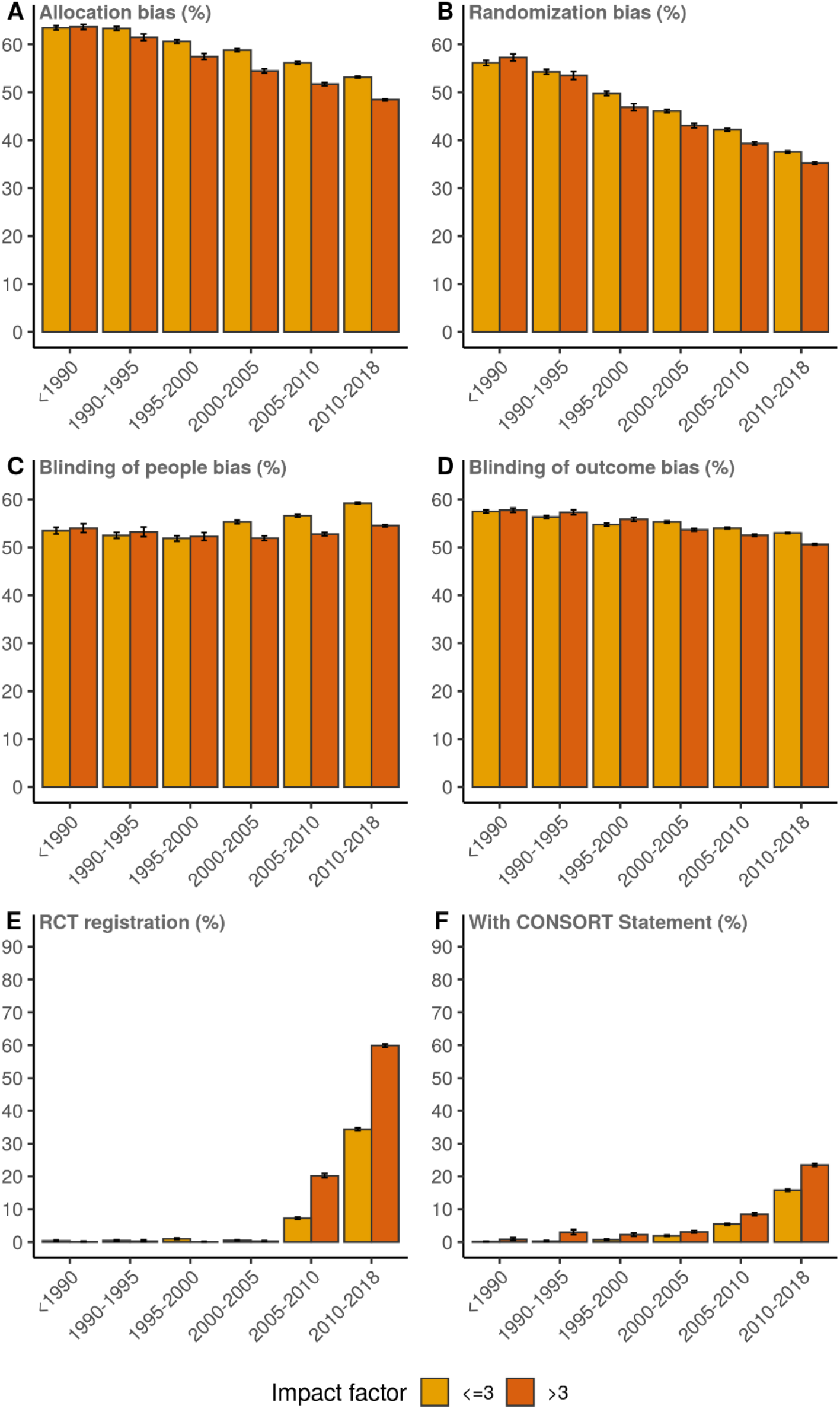
Risk of bias due to inadequate allocation concealment (A), random sequence generation bias (B), bias in blinding of patients and personnel (people) (C), bias in blinding of outcome assessment (D), RCT registration (E), and mentioning of the CONSORT Statement (F) plotted over time for RCTs published in journals with JIF>3 and journals with JIF<3. Indicated stratum range is up to but not including the last year.

**Supplementary Figure S3.**
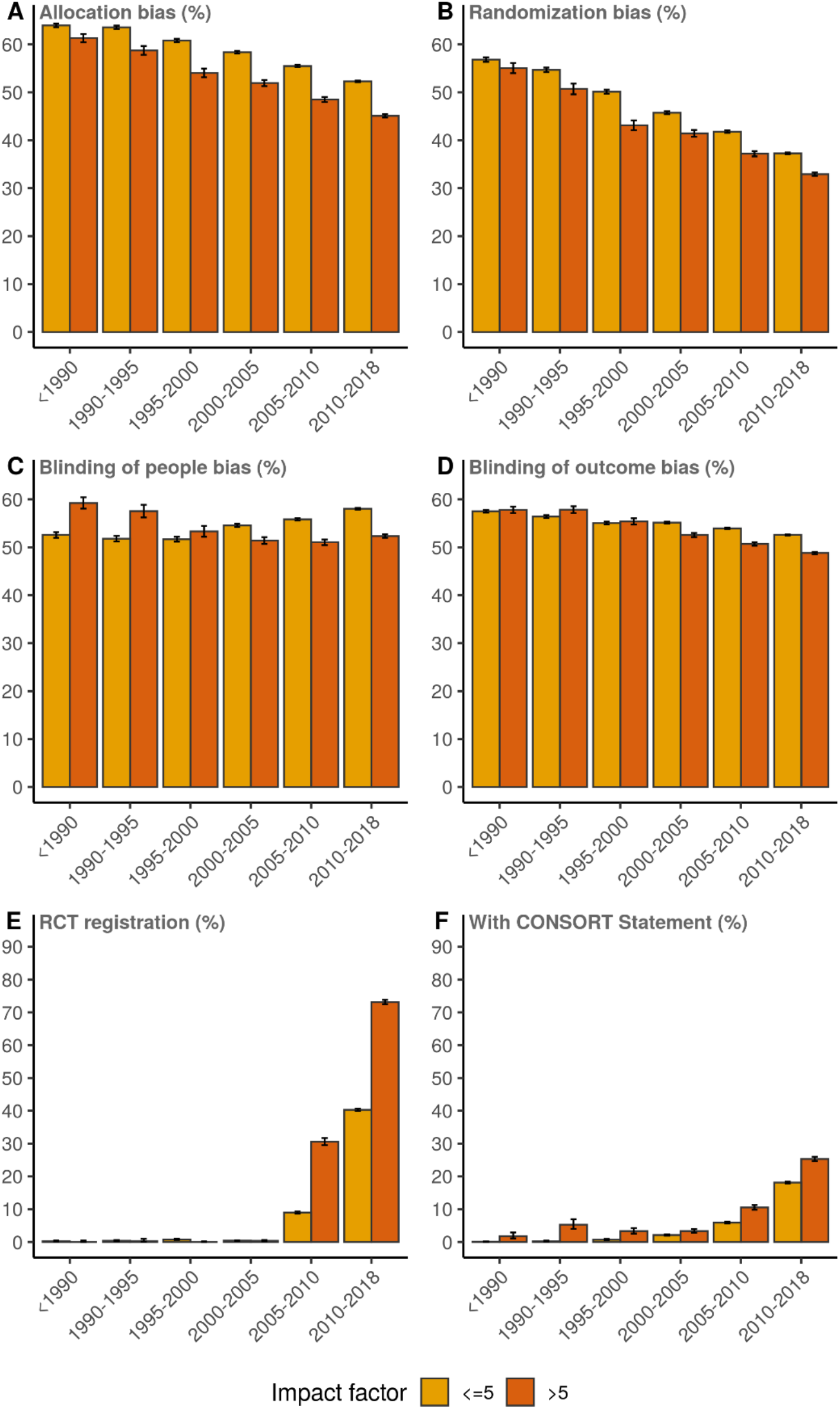
Risk of bias in allocation concealment (A), bias in randomization (B), bias in blinding of patients and personnel (people) (C), bias in blinding of outcome assessment (D), RCT registration (E) and mentioning of the CONSORT Statement (F) plotted over time for RCTs published in journals with JIF>5 and journals with JIF<5. Indicated stratum range is up to but not including the last year.

**Supplementary Figure S4.**
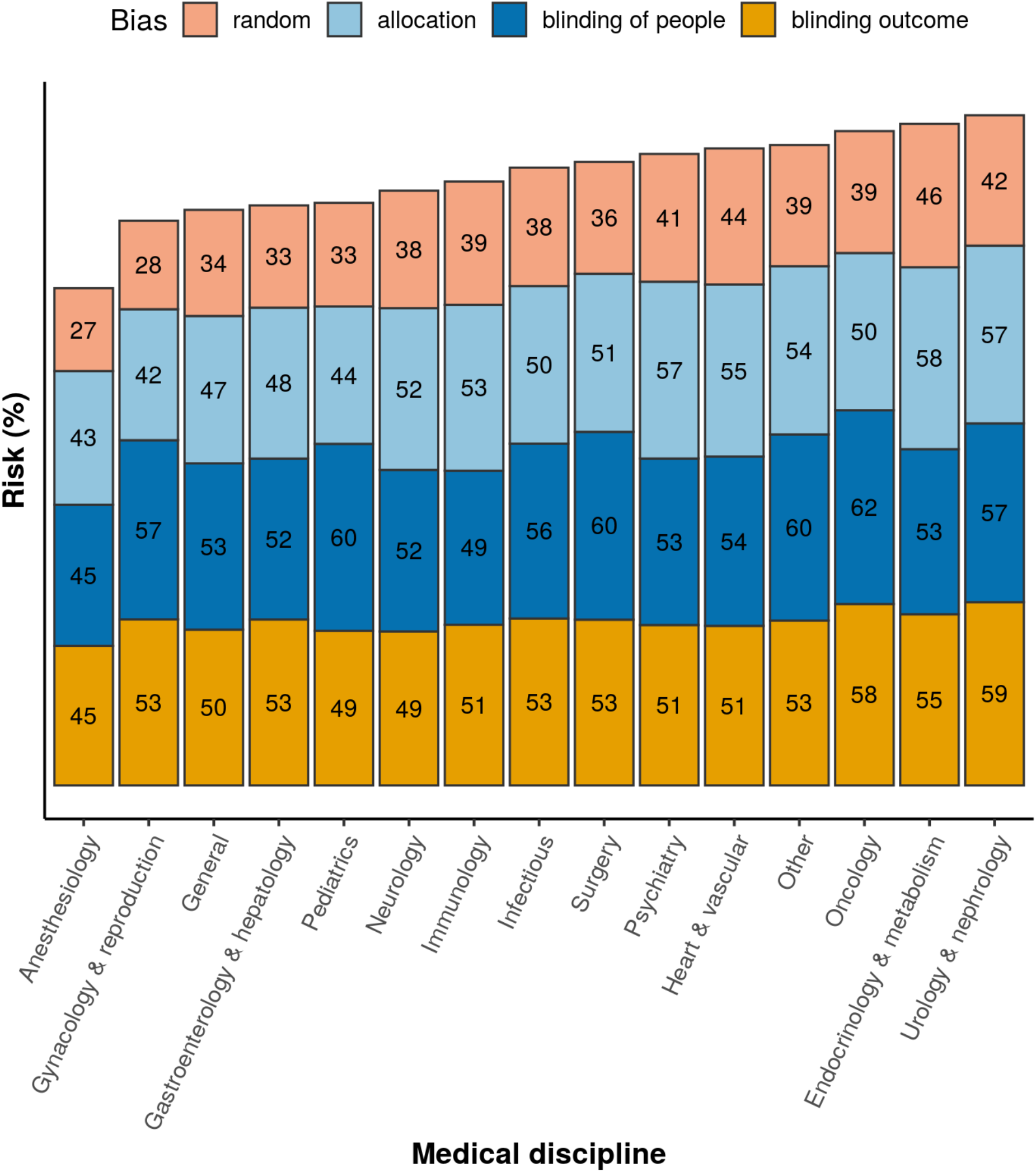
Average risk of biases, for trials published in the period 2005–2018 in different medical disciplines. ‘random’: bias in randomization; ‘allocation’: bias in allocation concealment; ‘blinding of people’: bias in blinding of patients and personnel; ‘blinding outcome’: bias in blinding of outcome assessment.

**Supplementary Figure S5.**
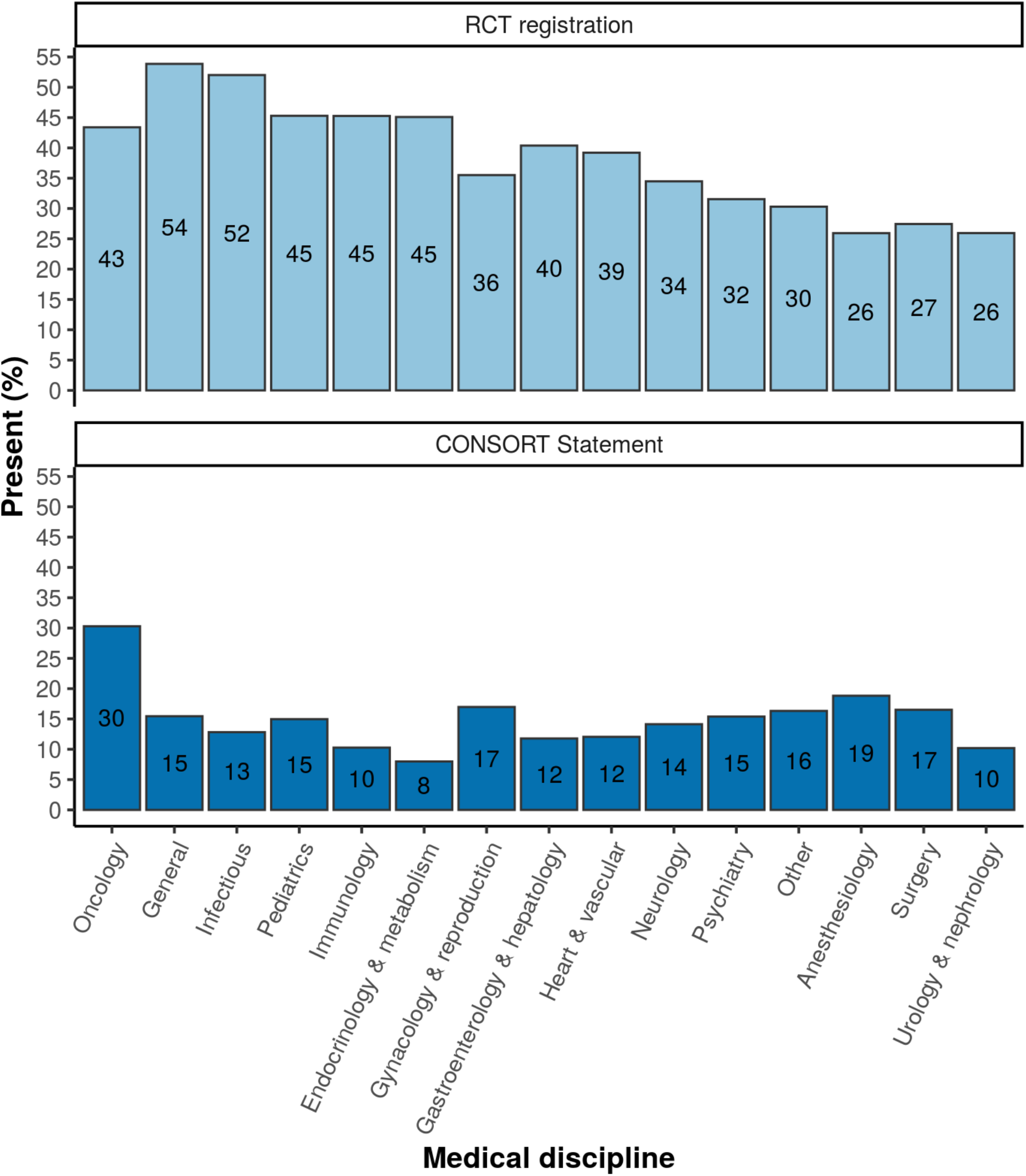
Presence of RCT registration and CONSORT Statement in trials published between 2005 and 2018.

**Supplementary Table S1.** Operationalization of variables for RCTs, authors, institutions, and journals.

| Variable | Details |
| --- | --- |
| **A. Randomized Clinical Trial** |  |
| Risk of Bias | Four domains of the Cochrane Risk of Bias tool: 1) random sequence generation, 2) allocation concealment, 3) blinding of participants and personnel, and 4) blinding of outcome assessment using validated machine learning approaches. Risk probabilities for the domains were extracted via open source software provided by RobotReviewer. RobotReviewer is developed to score risk of bias for four domains of the Cochrane Risk of Bias tool. No overall score was calculated. |
| Mention the CONSORT Statement in the publication | Yes/No |
| Mention of RCT registration in any public database (e.g. clinicaltrials.gov) | Yes/No |
| **B. Author** |  |
| Gender of first and last author | Based on the first name of an author, the API Genderize (https://genderize.io/) can determine the probability that this person is male or female. The gender with the highest probability was assigned to the author's name. |
| Proportion of female co-authors. | The putative gender of all authors was determined and combined into a proportion of all authors of the publication at issue. |
| Number of authors | Continuous number. |
| Number of countries | Total number of countries of (co-)authors. |
| Number of institutions | Total number of institutions of (co-)authors. |
| H(irsch)-index of first and last author | The H-index of the first and last author at time of publication was obtained from the Scopus web portal. |
| **C. Journal** |  |
| Medical discipline | Categories as downloaded from Web of Science inCites Journal Citation Reports, shortened list. If in multiple categories, the more specific category prevailed (e.g. cardiovascular vs. general medicine, or neurology vs. oncology). |
| Journal impact factor of the year before publication | JIF was extracted for each journal from Web of Science data for the period 1997–2016 as no earlier time points were available. The JIF's of 1997 were assigned to RCTs published before 1996. |

**Supplementary Table S1.** Total number (N) of trials published in the period 2005–2018 in the different medical disciplines with the number (K) and corresponding proportion (percentage with 95% confidence interval) of trials with a risk of bias probability below 50% (i.e. ‘low risk’).

| Medical discipline | Domain | N | K | Low risk (CI) |
| --- | --- | --- | --- | --- |
| **Anesthesiology** | Randomization | 5628 | 4482 | 79.6% (78.6-80.7%) |
|  | Allocation concealment |  | 3138 | 55.8% (54.4-57.1%) |
|  | Blinding of participants and personnel |  | 2984 | 53% (51.7-54.3%) |
|  | Blinding of outcome assessment |  | 3280 | 58.3% (57-59.6%) |
| **Endocrinology & metabolism** | Randomization | 6080 | 2861 | 47.1% (45.8-48.3%) |
|  | Allocation concealment |  | 1634 | 26.9% (25.8-28%) |
|  | Blinding of participants and personnel |  | 2393 | 39.4% (38.1-40.6%) |
|  | Blinding of outcome assessment |  | 1818 | 29.9% (28.8-31.1%) |
| **Gastroenterology & hepatology** | Randomization | 3751 | 2572 | 68.6% (67.1-70%) |
|  | Allocation concealment |  | 1704 | 45.4% (43.8-47%) |
|  | Blinding of participants and personnel |  | 1468 | 39.1% (37.6-40.7%) |
|  | Blinding of outcome assessment |  | 1345 | 35.9% (34.3-37.4%) |
| **General** | Randomization | 11907 | 7897 | 66.3% (65.5-67.2%) |
|  | Allocation concealment |  | 5618 | 47.2% (46.3-48.1%) |
|  | Blinding of participants and personnel |  | 4479 | 37.6% (36.7-38.5%) |
|  | Blinding of outcome assessment |  | 5170 | 43.4% (42.5-44.3%) |
| **Gynecology & reproduction** | Randomization | 5129 | 3870 | 75.5% (74.2-76.6%) |
|  | Allocation concealment |  | 2742 | 53.5% (52.1-54.8%) |
|  | Blinding of participants and personnel |  | 1554 | 30.3% (29-31.6%) |
|  | Blinding of outcome assessment |  | 1812 | 35.3% (34-36.7%) |
| **Heart & vascular** | Randomization | 9218 | 4902 | 53.2% (52.2-54.2%) |
|  | Allocation concealment |  | 3050 | 33.1% (32.1-34.1%) |
|  | Blinding of participants and personnel |  | 3360 | 36.5% (35.5-37.4%) |
|  | Blinding of outcome assessment |  | 3828 | 41.5% (40.5-42.5%) |
| **Immunology** | Randomization | 3638 | 2140 | 58.8% (57.2-60.4%) |
|  | Allocation concealment |  | 1278 | 35.1% (33.6-36.7%) |
|  | Blinding of participants and personnel |  | 1666 | 45.8% (44.2-47.4%) |
|  | Blinding of outcome assessment |  | 1470 | 40.4% (38.8-42%) |
| **Infectious** | Randomization | 3931 | 2471 | 62.9% (61.3-64.4%) |
|  | Allocation concealment |  | 1600 | 40.7% (39.2-42.3%) |
|  | Blinding of participants and personnel |  | 1276 | 32.5% (31-34%) |
|  | Blinding of outcome assessment |  | 1316 | 33.5% (32-35%) |
| **Neurology** | Randomization | 5269 | 3257 | 61.8% (60.5-63.1%) |
|  | Allocation concealment |  | 1977 | 37.5% (36.2-38.8%) |
|  | Blinding of participants and personnel |  | 2109 | 40% (38.7-41.4%) |
|  | Blinding of outcome assessment |  | 2330 | 44.2% (42.9-45.6%) |
| **Oncology** | Randomization | 6980 | 4392 | 62.9% (61.8-64.1%) |
|  | Allocation concealment |  | 2947 | 42.2% (41.1-43.4%) |
|  | Blinding of participants and personnel |  | 1535 | 22% (21-23%) |
|  | Blinding of outcome assessment |  | 1575 | 22.6% (21.6-23.6%) |
| **Other** | Randomization | 43051 | 25954 | 60.3% (59.8-60.7%) |
|  | Allocation concealment |  | 14775 | 34.3% (33.9-34.8%) |
|  | Blinding of participants and personnel |  | 11507 | 26.7% (26.3-27.2%) |
|  | Blinding of outcome assessment |  | 15432 | 35.8% (35.4-36.3%) |
| **Pediatrics** | Randomization | 2819 | 1945 | 69% (67.2-70.7%) |
|  | Allocation concealment |  | 1474 | 52.3% (50.4-54.1%) |
|  | Blinding of participants and personnel |  | 737 | 26.1% (24.5-27.8%) |
|  | Blinding of outcome assessment |  | 1234 | 43.8% (41.9-45.6%) |
| **Psychiatry** | Randomization | 7429 | 4277 | 57.6% (56.4-58.7%) |
|  | Allocation concealment |  | 2155 | 29% (28-30.1%) |
|  | Blinding of participants and personnel |  | 2986 | 40.2% (39.1-41.3%) |
|  | Blinding of outcome assessment |  | 3017 | 40.6% (39.5-41.7%) |
| **Surgery** | Randomization | 6912 | 4584 | 66.3% (65.2-67.4%) |
|  | Allocation concealment |  | 2792 | 40.4% (39.2-41.6%) |
|  | Blinding of participants and personnel |  | 1705 | 24.7% (23.7-25.7%) |
|  | Blinding of outcome assessment |  | 2455 | 35.5% (34.4-36.7%) |
| **Urology & nephrology** | Randomization | 3460 | 1935 | 55.9% (54.3-57.6%) |
|  | Allocation concealment |  | 996 | 28.8% (27.3-30.3%) |
|  | Blinding of participants and personnel |  | 1091 | 31.5% (30-33.1%) |
|  | Blinding of outcome assessment |  | 742 | 21.4% (20.1-22.9%) |

**Supplementary Table S2:**
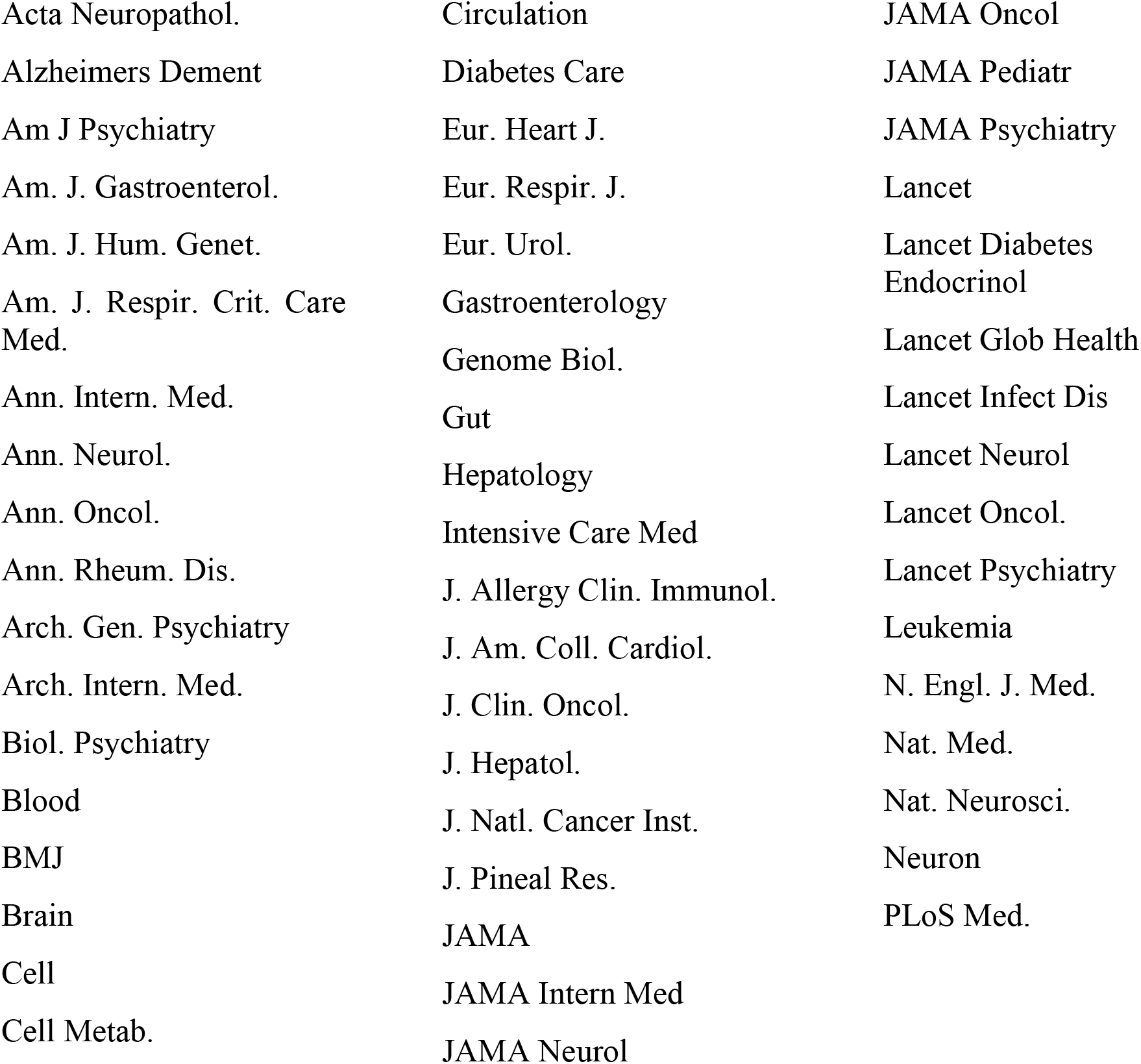
All journals with Journal Impact Factor (JIF) higher than 10 in year preceding any of the individual publications in our data set.

## References

1. Macleod MR, Michie S, Roberts I, et al. Biomedical research: increasing value, reducing waste. Lancet 2014;383(9912):101–4.

2. Prayle AP, Hurley MN, Smyth AR. Compliance with mandatory reporting of clinical trial results on ClinicalTrials.gov: cross sectional study. BMJ 2012;344:d7373.

3. Ioannidis JP. Why most discovered true associations are inflated. Epidemiology 2008;19(5):640–8.

4. Yordanov Y, Dechartres A, Porcher R, Boutron I, Altman DG, Ravaud P. Avoidable waste of research related to inadequate methods in clinical trials. BMJ 2015;350:h809.

5. Glasziou P, Altman DG, Bossuyt P, et al. Reducing waste from incomplete or unusable reports of biomedical research. Lancet 2014;383(9913):267–76.

6. Chalmers I, Glasziou P. Avoidable waste in the production and reporting of research evidence. Lancet 2009;374(9683):86–9.

7. Zwierzyna M, Davies M, Hingorani AD, Hunter J. Clinical trial design and dissemination: comprehensive analysis of clinicaltrials.gov and PubMed data since 2005. BMJ 2018;361:k2130.

8. Schulz KF, Altman DG, Moher D, Group C. CONSORT 2010 statement: updated guidelines for reporting parallel group randomised trials. BMJ 2010;340:c332.

9. Chalmers I, Bracken MB, Djulbegovic B, et al. How to increase value and reduce waste when research priorities are set. Lancet 2014;383(9912):156–65.

10. Powell-Smith A, Goldacre B. The TrialsTracker: Automated ongoing monitoring of failure to share clinical trial results by all major companies and research institutions. F1000Res 2016;5:2629.

11. Moher D, Bouter L, Kleinert S, et al. The Hong Kong Principles for Assessing Researchers: Fostering Research Integrity. OSF Preprints 2019;17 Sept 2019.

12. Turner L, Shamseer L, Altman DG, et al. Consolidated standards of reporting trials (CONSORT) and the completeness of reporting of randomised controlled trials (RCTs) published in medical journals. Cochrane Database Syst Rev 2012;11:MR000030.

13. Dechartres A, Trinquart L, Atal I, et al. Evolution of poor reporting and inadequate methods over time in 20 920 randomised controlled trials included in Cochrane reviews: research on research study. BMJ 2017;357:j2490.

14. Damen JA, Lamberink HJ, Tijdink JK, et al. Predicting questionable research practices in randomized clinical trials. Open Science Framework, https://osf.io/27f53/ 2018;December 19th, 2018.

15. Vinkers CH, Tijdink JK, Otte WM. Use of positive and negative words in scientific PubMed abstracts between 1974 and 2014: retrospective analysis. BMJ 2015;351:h6467.

16. Millard LA, Flach PA, Higgins JP. Machine learning to assist risk-of-bias assessments in systematic reviews. Int J Epidemiol 2016;45(1):266–77.

17. Marshall IJ, Kuiper J, Wallace BC. RobotReviewer: evaluation of a system for automatically assessing bias in clinical trials. J Am Med Inform Assoc 2016;23(1):193–201.

18. Gates A, Vandermeer B, Hartling L. Technology-assisted risk of bias assessment in systematic reviews: a prospective cross-sectional evaluation of the RobotReviewer machine learning tool. J Clin Epidemiol 2018;96:54–62.

19. Higgins JP, Altman DG, Gotzsche PC, et al. The Cochrane Collaboration’s tool for assessing risk of bias in randomised trials. BMJ 2011;343:d5928.

20. Viergever RF, Li K. Trends in global clinical trial registration: an analysis of numbers of registered clinical trials in different parts of the world from 2004 to 2013. BMJ Open 2015;5(9):e008932.

21. Garfield E. The history and meaning of the journal impact factor. JAMA 2006;295(1):90–3.

22. West JD, Jacquet J, King MM, Correll SJ, Bergstrom CT. The role of gender in scholarly authorship. PLoS One 2013;8(7):e66212.

23. De Angelis CD, Drazen JM, Frizelle FA, et al. Is this clinical trial fully registered? A statement from the International Committee of Medical Journal Editors. Lancet 2005;365(9474):1827–9.

24. Mathieu S, Boutron I, Moher D, Altman DG, Ravaud P. Comparison of registered and published primary outcomes in randomized controlled trials. JAMA 2009;302(9):977–84.

25. Lo B. Sharing clinical trial data: maximizing benefits, minimizing risk. JAMA 2015;313(8):793–4.

26. Gluud LL, Sorensen TI, Gotzsche PC, Gluud C. The journal impact factor as a predictor of trial quality and outcomes: cohort study of hepatobiliary randomized clinical trials. Am J Gastroenterol 2005;100(11):2431–5.

27. Konsgen N, Barcot O, Hess S, et al. Inter-review agreement of risk-of-bias judgments varied in Cochrane reviews. J Clin Epidemiol 2019;120:25–32.

28. Armijo-Olivo S, Ospina M, da Costa BR, et al. Poor reliability between Cochrane reviewers and blinded external reviewers when applying the Cochrane risk of bias tool in physical therapy trials. PLoS One 2014;9(5):e96920.

29. Marshall IJ, Kuiper J, Banner E, Wallace BC. Automating Biomedical Evidence Synthesis: RobotReviewer. Proc Conf Assoc Comput Linguist Meet 2017;2017:7–12.

30. Begg C, Cho M, Eastwood S, et al. Improving the quality of reporting of randomized controlled trials. The CONSORT statement. JAMA 1996;276(8):637–9.

